# Impact of Personal Care Habits on Post-Lockdown COVID-19 Contagion: Insights from Agent-based Simulations

**DOI:** 10.1101/2020.09.23.20200212

**Authors:** Lindsay Álvarez-Pomar, Sergio Rojas-Galeano

## Abstract

After the first wave of spread of the COVID-19 pandemic, countries around the world are struggling to recover their economies by slowly lifting the mobility restrictions and social distance measures enforced during the crisis. Therefore, the post-lockdown containment of the disease will depend strongly not any more on government-imposed interventions but on personal care measures, taken voluntarily by their citizens. In this respect, recent studies have shed some light regarding the effectiveness individual protection habits may have in preventing SARS-Cov-2 transmission, particularly physical contact distancing, facial mask wearing and hand-washing habits. In this paper we describe experiments performed on a simulated COVID-19 epidemic in an artificial population using an agent based model, so as to illustrate to what extent the interplay between such personal care habits contributes to mitigate the spread of the disease, assuming the lack of other population-wide non-pharmaceutical interventions or vaccines. We discuss scenarios where wide adherence to these voluntary care habits alone, can be enough to contain the unfold of the contagion. Our model purpose is illustrative and contributes to ratify the importance of disseminating the message regarding the collective benefits of mass adoption of personal protection and hygiene habits, as an exit strategy for COVID-19 in the new normal state.

## Introduction

Countries around the world are facing the extraordinary challenge of recovering the economy after the crisis brought about by the COVID-19 pandemic (***Pichler et al., 2020; Fernandes, 2020; Coibion et al., 2020***). The crisis yielded negative impact on social, economic and psychological conditions of the population due to the application of Non-Pharmaceutical Interventions (NPI) intended to limit the mobility of people, reducing in this way the risk of contagion by direct contact, e.g. total lock-down, home quarantines, isolation of confirmed cases, closing conglomeration facilities (***Ferguson et al., 2020; Lai et al., 2020; Giordano et al., 2020; Kantner, 2020***).

After a first peak of infections, most countries have decided to lift these kind of NPIs in order to reopen their economies, allowing susceptible persons to move around freely, despite the continuing prevalence of the virus and the lack of approved vaccines (to this date). Thus, the major risk of contagion would be caused by sustained person-to-person contact with asymptomatic SARS-CoV-2 patients, particularly in crowded gatherings or poorly ventilated spaces (***Harris, 2020; Allen and Marr, 2020; Qian et al., 2020***). Consequently, individual protection measures taken voluntarily by the citizens in order to minimise such person-to-person contact risk (e.g. maintaining physical proximity distance, wearing face protection equipment and washing hands regularly), would be of extreme importance to combat a resurgence of COVID-19 transmission in the new reopening scenario. In other words, they will play a key role in preventing not only the spread of the disease but also the need of sending communities back into lockdown.

Compared to the socioeconomic cost inflicted by government-mandate NPIs, it may be argued that personal protection measures are seemingly inexpensive (or at least simple to deploy) and yet, highly effective in preventing SARS-CoV-2 contagion (***Chu et al., 2020***). A study showed that washing hands with a frequency of 6 to 10 times per day yields a reduction of personal risk of infection of about 34%, compared to individuals that washed them with lower frequency (***Beale et al., 2020***). Other studies have previously linked hand hygiene as an effective countermeasure against similar respiratory infectious diseases such as influenza (***Liu et al., 2016; Abdulrahman et al., 2019***).

In a similar vein, given that airborne transmission has been identified as the principal route for the spread of SARS-CoV-2 (***Zhang et al., 2020***), the habit of wearing facial mask protection that once was dismissed as innocuous by the World Health Organization (***WHO, 2020***), now it is being recommended as a necessary barrier to filter the enveloped droplet-borne shedding mechanism of this respiratory virus (***Leung et al., 2020; Zhang et al., 2020***).

Reducing outward shedding and thus, reducing contamination of the environment are the major benefits of adopting this habit. The beneficial filtering effect both inwards and outwards of masks made out of different cloth materials has been documented (***Clase et al., 2020***). Some studies found a reduced risk of getting infected for healthy mask users, but in addition, if anyway infection occurs, masks reduced the amount of virus particles the susceptible person was exposed to, presumably leading to a mild or asymptomatic infection (***Gandhi and Rutherford, 2020***). As a result, universal masking could become an alternative mechanism to community-level immunisation, before a safe and approved vaccine arrives (***Gandhi et al., 2020***).

Another observational study reported successful protection of a community using facial masks, while attending a hair salon where two stylists (who also wore facial protection) were later on diagnosed with COVID-19 and developed symptoms; none of these clients become infected with the disease (***Hendrix, 2020***). A retrospective study of secondary infections in households revealed that mask wearing by the index case and family contacts before onset of the patient’s symptoms, yielded a significant reduction in the risk of transmission (***Wang et al., 2020***).

Although no sufficient evidence is yet available to estimate the exact ratio of risk reduction provided by face mask protection, an overall trend of epidemic mitigation has been noticed in several countries after this measure is adopted by the majority of the community (***Howard et al., 2020; Mitze et al., 2020***). Similar studies regarding combination of measures involving physical contact distancing, face mask and eye protection hint at their efficacy to prevent person-to-person transmission of SARS-CoV-2 (***Chu et al., 2020***). It has been suggested that in community contexts, masks appeared to be effective with and without hand hygiene, or even more protective if both together are taken (***MacIntyre and Chughtai, 2020***).

In view of this background, our study addresses the following question: To what extent personal protection habits can be effective in containing the spread of the post-lockdown COVID-19 contagion, in absence of any other NPIs or treatment, when examined within a controlled simulated environment? As our findings indicate indeed, mitigation effects emerge when the majority of the population adheres to these protection measures. Our computational model purpose is illustrative (***Squazzoni et al., 2020***), and contributes to ratify the importance of reaching out across the community to disseminate the message about the collective benefits of mass adoption of personal care against COVID-19, under the new normal state.

## Methods and tools

We build upon the agent-based simulation of COVID-19 epidemic introduced in (***Alvarez and Rojas-Galeano, 2020***). That model was designed to simulate the dynamics of the COVID-19 contagion within a population of a toy city, based-on a SIRE+CARDS epidemic model considering four states: Susceptible, Infectious, Recovered and Extinct. The Infectious is actually seen as a macro-stateincluding conditions Confirmed, Asymptomatic, Risky, Deadly and Severe. The contagion unfolds as infectious agents go out of their households and wonder around the city while interacting with other healthy agents. In addition, a number of NPIs are available including lockdowns, quarantines and mass-testings. For more details of the agent-based design, the epidemic model, the transition events, and the NPIs dynamics, we refer the reader to (***Alvarez and Rojas-Galeano, 2020***).

We proceeded to extend the model to account for individual voluntary protection measures, related to personal health habits, namely physical distance, facial masks wearing and regular hand-washing. Consequently, each agent was designed with three different personal habit traits: social-distancer (SD), mask-user (MU) and hand-washer (HW). The actual choice of traits for an arbitrary agent at the beginning of the simulation, is controlled with three adjustable parameters that represent the willingness of the total population to adopt them, in a proportion between 0 and 100%.

The incidence of these personal habits in the risk of contagion during person-to-person contact was modelled as follows. Firstly, for social-distancer agents the chance of a direct contact will be diminished as a result of their tendency to divert their trajectory to avoid a close encounter. The actual occurrence of contact events would depend on the random spatial interactions agents will have as they move around the city during their daily activities, as the simulation unfolds.

Secondly, after pondering the evidence favouring the efficacy of facial masks to prevent SARS-CoV-2 transmission (***Wang et al., 2020; Leung et al., 2020; Mitze et al., 2020; Chu et al., 2020; Gandhi et al., 2020; Clase et al., 2020; Zhang et al., 2020; Hendrix, 2020; Howard et al., 2020***), we defined four possible cases of protection configuration during a single contagion event involving one susceptible and one infectious agents, along with their associated risks of transmission: if none of the agents are wearing masks, the chance of contagion would be 90%; if susceptible wears mask but infectious does not, the chance of contagion would be 50%; if infectious wears mask but susceptible does not, the chance of contagion would be 30%; the last case correspond to both agents wearing masks, with a chance of contagion of 10%.

Thirdly, and again taking into account the recent studies suggesting the benefits of the hand-washing habit in preventing coronavirus-like diseases (***Beale et al., 2020; Liu et al., 2016***), we defined a further reduction of the contagion risk in a 30% factor if the susceptible agent involved in any of the above described contagion events happens to be a hand washer (e.g, in the case where infectious agent wears masks but susceptible does not, the risk decreases from 30% to 9%).

Given that the purpose of our study is to assess the effect these habits can have in the mitigation of the epidemic, we decided to call off the application of any of the other NPIs available in the model, and experiment with simulations where the population to some degree adheres only to SD, MU and HW habits. For each of these traits, we defined willingness parameters modelling scenarios where nobody observes the habit (0%), approximately half of the population adopt it (50%) or everybody adheres to it (100%). Thus, twenty-seven scenarios were considered corresponding to different permutations of the tuple of proportions (SD%, MU%, HW%), beginning with the organic “do nothing” scenario (0%, 0%, 0%) up to the ideal “everybody adheres to” scenario (100%, 100%, 100%) plus the in between permutations: (0%, 0%, 50%), (0%, 0%, 100%), (0%, 50%, 0%), etc.

Besides, our analysis will consider the following epidemic indicators:

1. *Mortality*. This measure is the cumulative count of deaths in the population. Notice that in our model all deaths are due to COVID-19, and no births or immigration are taking into account during the simulation timeline. Thus, this indicator determines the mortality rate.
2. *Cases*. This measure is the cumulative count of infections in the population. Since the population evolves within a controlled environment where both symptomatic and asymptomatic patients can be traced, this would be in fact the actual number of cases due to the disease. This indicator is associated to the Infection Fatality Rate (IFR).
3. *Conirmed cases*. This measure is the cumulative count of confirmed infection cases. Since in our simulations we did not enable the application of mass-testing interventions, these cases correspond to symptomatic patients whose disease worsens to severe or deadly states requiring hospital care, where upon admission, are reported as diagnosed. This indicator is associated to the Case Fatality Rate (CFR) which is usually an overestimated representative of incidence rate (i.e, CFR ≫ IFR).
4. *Recovered*. This measure is the cumulative count of agents that recovered from disease. The model assumes that upon recovery. immunity to the disease is acquired and no re-infection will occur. Therefore, this indicator is related to the survival rate and herd immunity rate.

The derived model was developed in the NetLogo programming language version 6.1.0; the source-code and user documentation have been released openly in: http://modelingcommons.org/browse/one_model/6423. There, the model can be run online or downloaded for local execution.

## Simulation results

The setup of the experiments, including settings for the artificial city, population, disease, NPIs and running parameters are shown in Table 1 (notice that for the sake of completeness, we report settings for authority-enforced NPIs although they were disabled during the simulations). A single simulation starts at 00 hours of day 0, and runs until 00 hours of day 60. At 12h of day 0 an outbreak is simulated causing 5% of the population to get infected. From that moment, the simulation unfolds according to the rules designed for the model, and to the random local interactions occurring during the movement of the agents.

**Table 1.** Settings used in the simulation tool to perform the experiments.

| Parameter | Description | Value |
| --- | --- | --- |
| *City and population settings* |  |  |
| pop-size | Total number of simulated people (agents) | 400 |
| zones | Number of residential zones | 9 |
| days | Period of observation days (simulation length) | 60 days |
| % high-risk | % of population with co-morbidities | 30% |
| hospital-beds | Total number of hospital beds available | 12 |
| ICU-beds | Total number of ICU beds available | 2 |
| ambulances-zone | Number of ambulances (sentinels) per zone | 1 |
| *Disease settings* |  |  |
| avg-duration | Average day period to recover from illness | 18 days |
| % asymptomatic | % of patients showing no or mild symptoms | 50% |
| *NPI settings (authority-enforced, not used)* |  |  |
| total-lockdown? | Enforce stay-at-home order for the entire population | off |
| % permits | % of mobility permits when lockdown is activated | not used |
| case-isolation? | Confirmed cases are isolated at home | off |
| home-quarantine? | Housemates of confirmed cases are also isolated | off |
| zone-enforcing? | Restraint mobility of agents within zones only | off |
| pick-zone | Zone id number | not used |
| sentinel-testing? | Enable mass-testing by ambulance sentinels | off |
| zonal? | Restraint mobility of sentinels within zones | off |
| *NPI settings (personal-habits)* |  |  |
| social-distancing? | Maintain a minimum physical distance with others (SD) | on |
| % willings | Estimated % of population willing to comply with SD | {0, 50, 100} |
| % mask-users | Estimated % of population willing to use masks (MU) | {0, 50, 100} |
| % hand-washers | Estimated % of population willing to wash hands (HW) | {0, 50, 100} |

### Effect in “lattening the curve”

An illustration of the SIRE curves obtained in a single representative run for each of the 27 scenarios are shown in Figures 1, 2 and 3. Firstly, Figure 1 reveals that most of the plots when no social distancing is adopted (SD=0%), exhibit the typical unfold of an uncontrolled epidemic, that is a exponential grow of the infectious curve (red) achieving an early peak in the first third of the simulation. Only the scenarios where all the agents wear mask protection (MU=100%) and half or more of the agent population wash hands regularly (HW=50%, 100%), the infectious curve developed a flattened shape indicating a mitigation on the speed of contagion.

**Figure 1.**
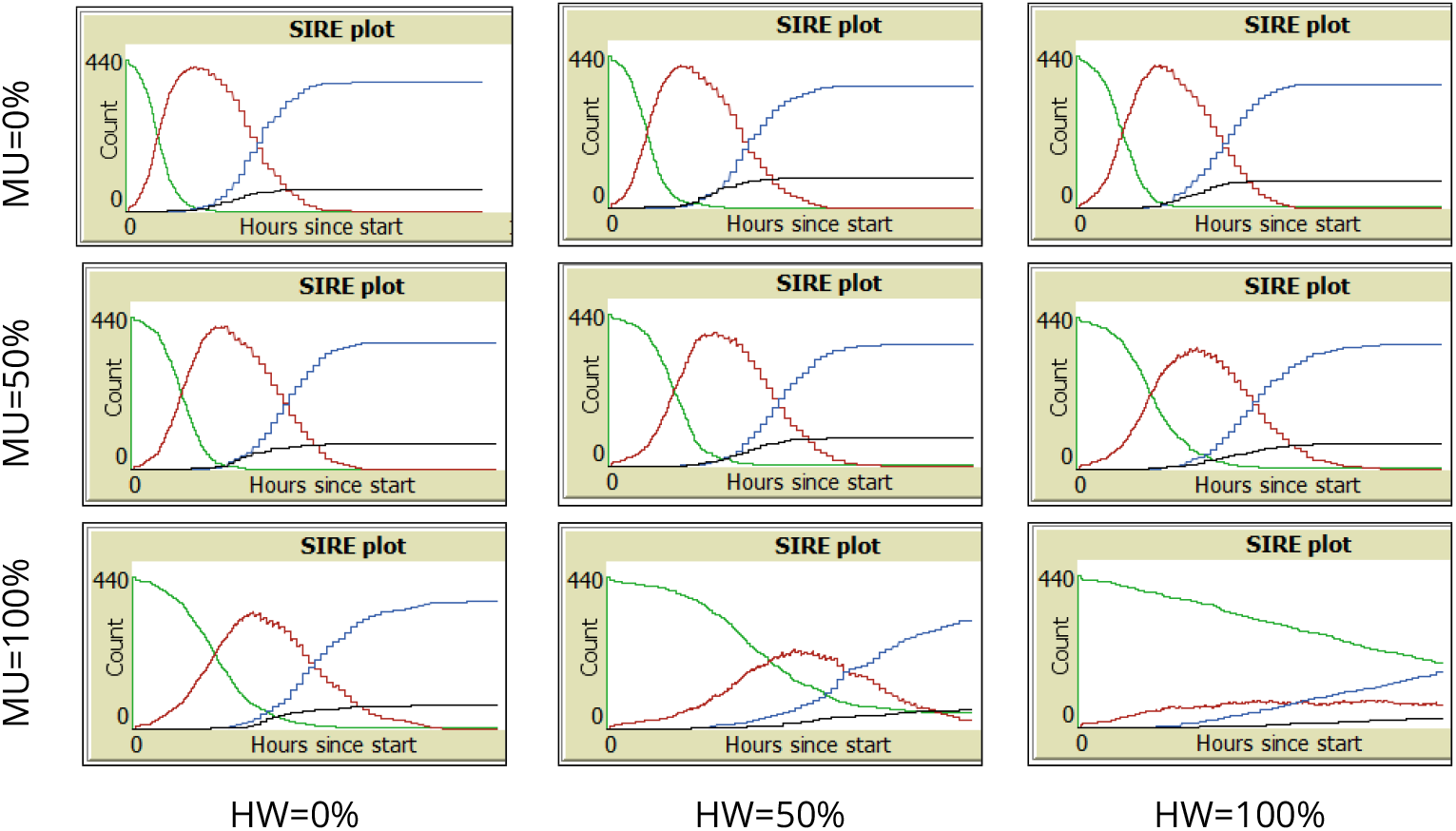
Representative SIRE curves for scenarios with SD=0% and different combinations of MU and HW proportions (green: Susceptible, red: Infectious, blue: Recovered, black: Extinct).

**Figure 2.**
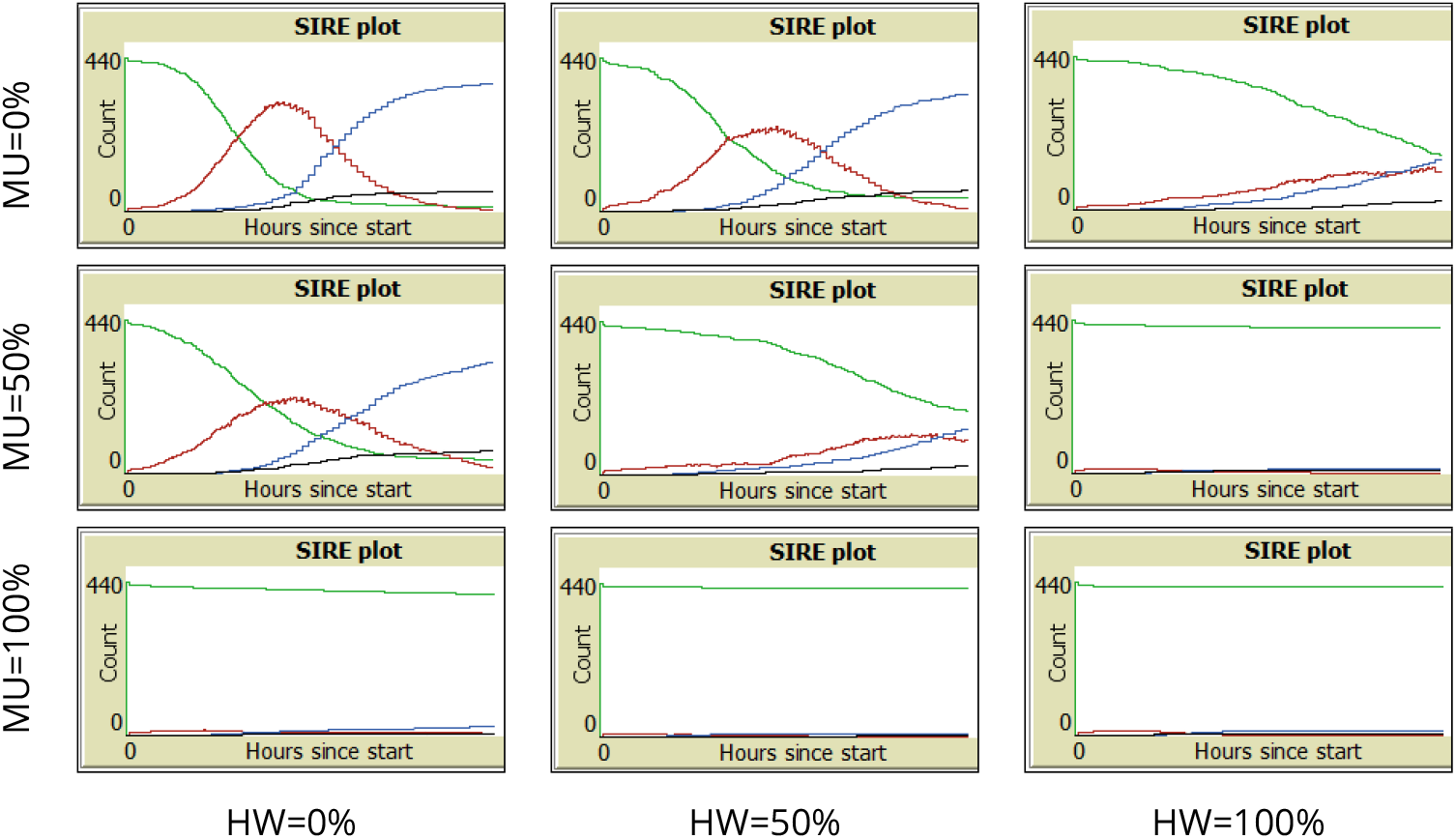
Representative SIRE curves for scenarios with SD=50% and different combinations of MU and HW proportions (green: Susceptible, red: Infectious, blue: Recovered, black: Extinct).

**Figure 3.**
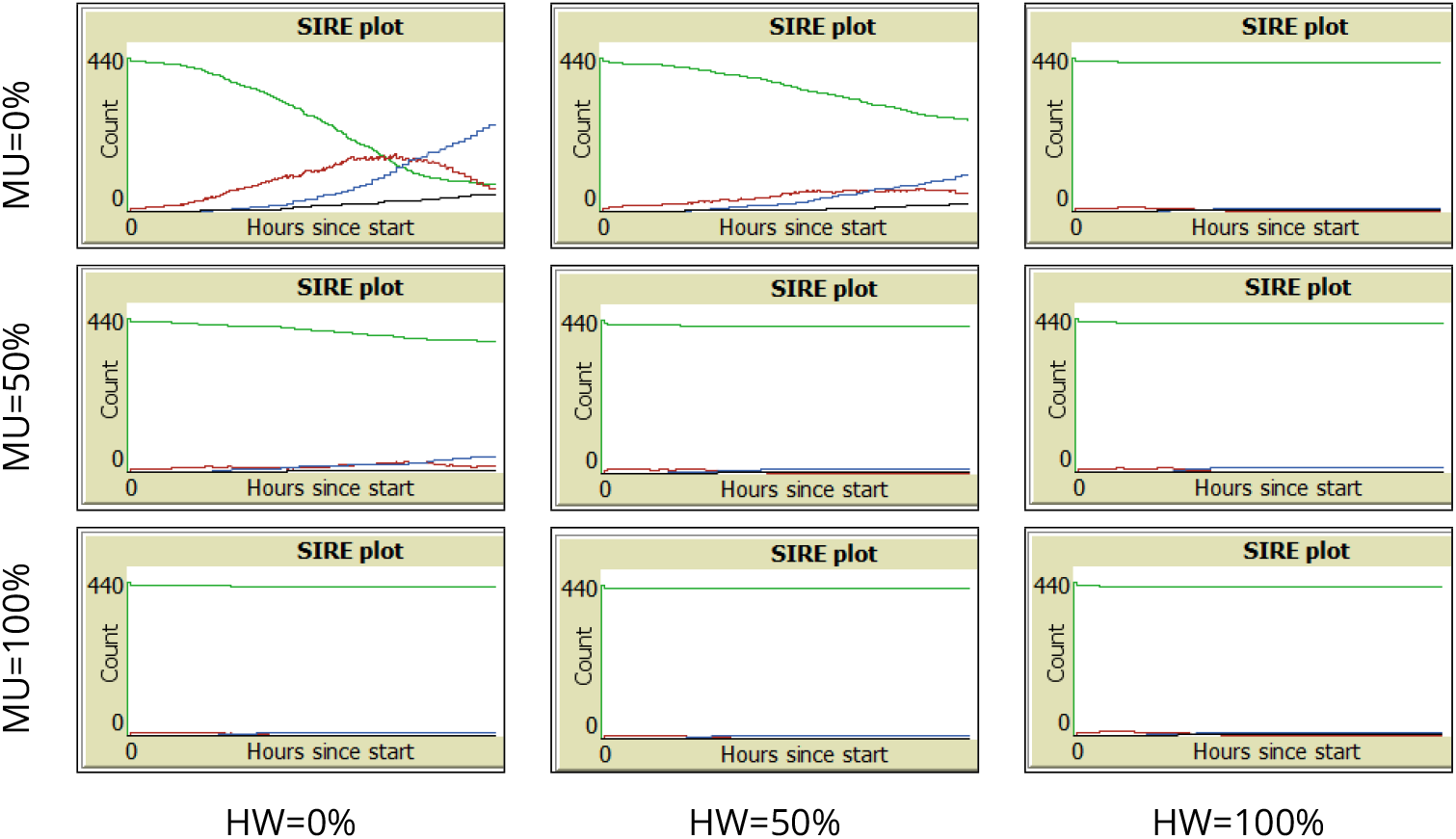
Representative SIRE curves for scenarios with SD=100% and different combinations of MU and HW proportions (green: Susceptible, red: Infectious, blue: Recovered, black: Extinct).

On the other hand, Figure 2 shows that a 50% increment of agents adhering to social distance (SD=50%) has a noticeable effect in the mitigation of the infectious curve. Even when no other habit is adopted (MU=0%, HW=0%) the peak is lowered and shifted towards the middle of the simulation timeline. When any other habit is adopted to some extent, the curve is flattened more drastically (MU=50% or HW=50%). Moreover, when more than 50% of the population adopts mask wearing or washing hands or both (MU=100%, HW=100%), the transmission of the infection is suppressed, even from the early stages of the simulation.

Lastly, Figure 3 shows that all scenarios where everybody adheres to maintain physical distance (SD=100%) and abide by the other habits to some extent (MU *>* 0%, HW *>* 0%), result in a remarkable effect of suppressing the contagion. Indeed, when nobody uses masks or washes hands (MU=0%, HW=0%), that is, only universal compliance with physical distance is observed, the infectious curve is clearly flattened and shifted towards the end of the simulation.

### Effect on the epidemic indicators

For each of the 27 SD-MU-HW scenarios we performed 30 repetitions. Then, at the end of the simulation run (day 60), we collected average statistics of the aforementioned epidemic indicators. We focused the analysis of these results, on assessing the effect of variations in the estimated ratios of population adopting personal care habits, that is, we compare the results of the epidemic indicators for a number of variations SD-MU-HW. For this purpose we defined as baseline three scenarios: in the first one, nobody adopts any habit (0%, 0%, 0%); in the second, half the population comply with physical distance but do not use masks or wash hands (50%, 0%, 0%); lastly, the third baseline scenario assumes everybody observe physical distance but again, nobody use masks or wash hands (100%, 0%, 0%). The differences in the behaviour between baseline scenarios and variations, were assessed using the Mann-Whitney U statistical test (see Supplementary Material).

Next we report the results for the epidemic indicators of interest. In the following plots, some periodic patterns can be seen when groups of three boxes are examined from left to right (a box with higher average, then a box in-between and then a lower average box) which are just an artefact of the order we chose to present the permutation of the tuple (SD, MU, HW), with each parameter taking values 0%, 50% and 100%. We remark that permutations in different order exhibit similar patterns with falling rates in their respective epidemic indicators (in cycles of every three scenarios).

### Mortality

The behaviour of this epidemic indicator across all the simulated scenarios is shown in Figure 4. The left panel shows scenarios where no one complies with physical distancing (SD=0%). No significant difference in the firsts 5 scenarios were found with respect to the (0%, 0%, 0%) baseline (number of deaths around 65, i.e 16% of the population). Only the last three scenarios in that panel, where everyone uses face masks (MU=100%), showed a significant decrease in the number of deaths, with the sharpest fall when also everyone washes hands (HW=100%) to around the half.

The middle panel shows scenarios where half of the population adheres to physical distancing (SD=50%). Within each group of 3 boxes, a trend can be seen where the number of deaths decreases as the percentage of HW increases. In any case, the number of deaths in all scenarios drops significantly compared to the baseline (50%, 0%, 0%) scenario for this panel. As expected, the lasts group of three scenarios obtained the lowest values, approximately fewer than 10 deaths (i.e 2.5% of the population).

**Figure 4.**
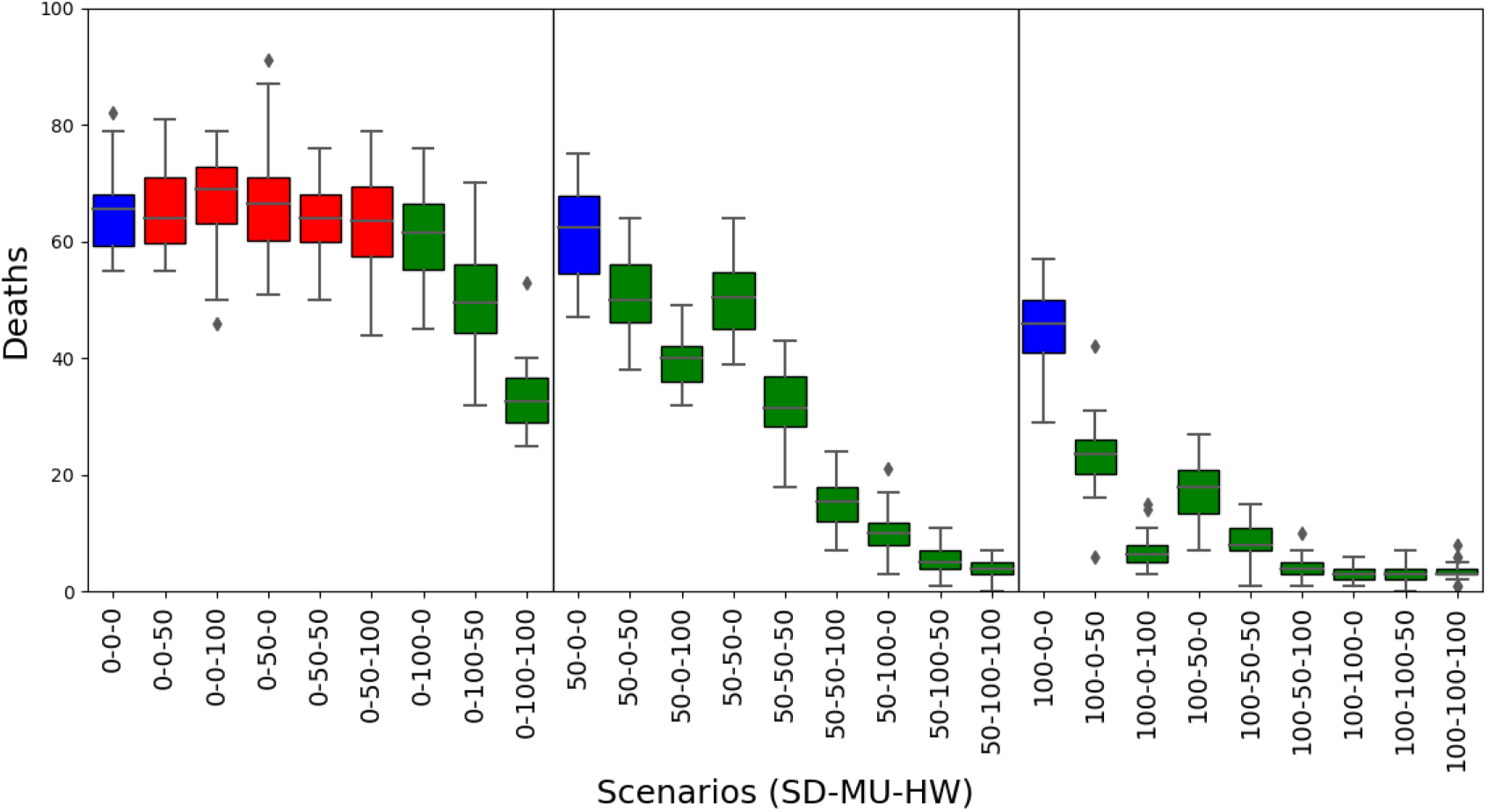
*Mortality* results. Box plots represent average and standard deviation of the cumulative count of deaths in each scenario. Scenarios are labelled according to the willingness of agents to adhere to personal health habits such as Social Distance, Mask User or Hands Washer (SD%-MU%-HW%). Baseline scenarios were defined varying the proportion of social distancers in the population assuming no agents adopt using masks or washing hands. In each panel, the baseline scenario is coloured blue (left: 0%-0%-0%, middle: 50%-0%-0%, right: 100%-0%-0%). Scenarios with statistical significant difference compared to their corresponding panel baseline are coloured green (*p*-value *<* 0.05), otherwise are coloured red.

The right panel shows scenarios where the entire population observes physical distancing (SD=100%). A similar pattern to the results of middle panel can be seen, as comparing to the baseline (100%, 0%, 0%) scenario, all the other scenarios exhibited a significant reduction in mortality, including the last six with death ratios fewer than 2.5% of the population. These results are corroborated with the black coloured Extinct curve of the SIRE plots of Figure 3, which is seen rising very low or even not rising at all.

### Cases

The behaviour of this epidemic indicator across all the simulated scenarios is shown in Figure 5. The three panels are organised as before: scenarios with SD=0% are plotted in the left panel, scenarios with SD=50% in the middle, and scenarios with SD=100% in the right panel. Regarding the left panel, a significant drop in the number of cases can be seen clearly on the (0%, 100%, 50%) and (0%, 100%, 100%) scenarios, around 350 and 270 cases, respectively; the difference is not noticeable with respect to the other scenarios where, the number of cases approaches the entire population (nearly 400 agents). In the middle panel a significant decrease in the number of cases can be seen for all scenarios. The largest drop is observed between the (50%, 50%, 50%) scenario and the (50%, 50%, 100%) scenario (around half of the cases, from 250 to 120 cases).

**Figure 5.**
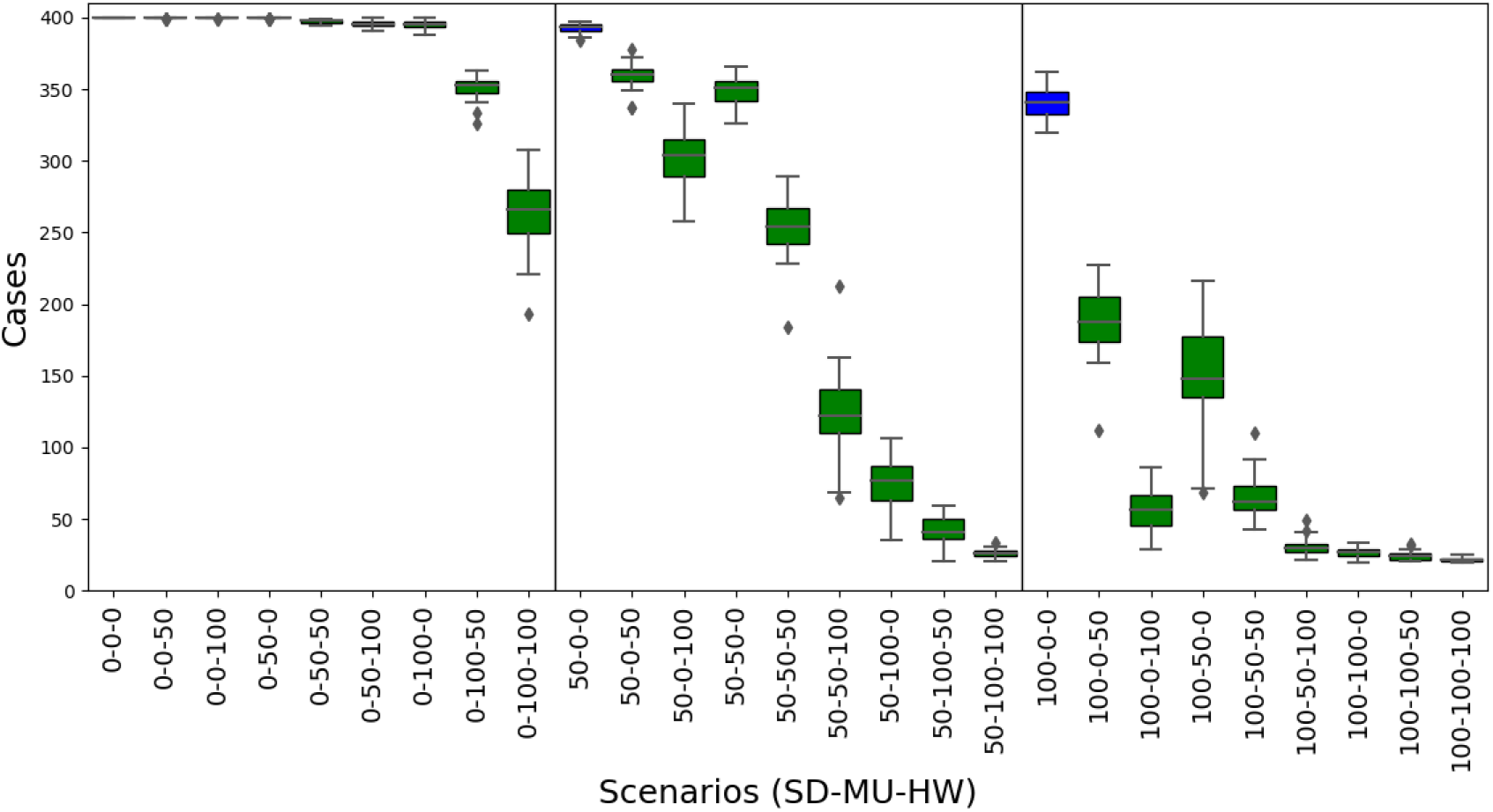
*Cases* results. Box plots represent average and standard deviation of the cumulative count of infections in the population. See caption of Fig. 4 for interpretation details.

Lastly, in the right panel, a similar pattern to the middle panel can be seen, although more notably: compared to the baseline (100%, 0%, 0%) for this panel, the other scenarios exhibited a significant reduction in cases, approaching around 25 (6.25% of the population) cases in the last four scenarios. These low rates are explained because, as Figure 2 and Figure 3 show, the red Infectious curve in some of these scenarios is flattened, and therefore the contagion is at the stage of initial growth when the indicator was collected at the end of the simulated timeline (day 60); what is more, in some other cases the curve was entirely suppressed indeed.

### Confirmed cases

The behaviour of this epidemic indicator across all the simulated scenarios is shown in Figure 6. A similar pattern with the mortality plots of Figure 4 can be seen in the three panels. In contrast to mortality, the value of confirmed cases is higher than deaths (notice the three baseline scenarios approaching around 90 confirmed cases in average). We remark that the confirmed cases results are much lower compared to the actual cases results, because in these simulations we did not applied mass testing, voluntary isolation or home quarantine, therefore diagnosed cases correspond only to those admitted (and reported as cases) to hospital due to complications with the disease.

**Figure 6.**
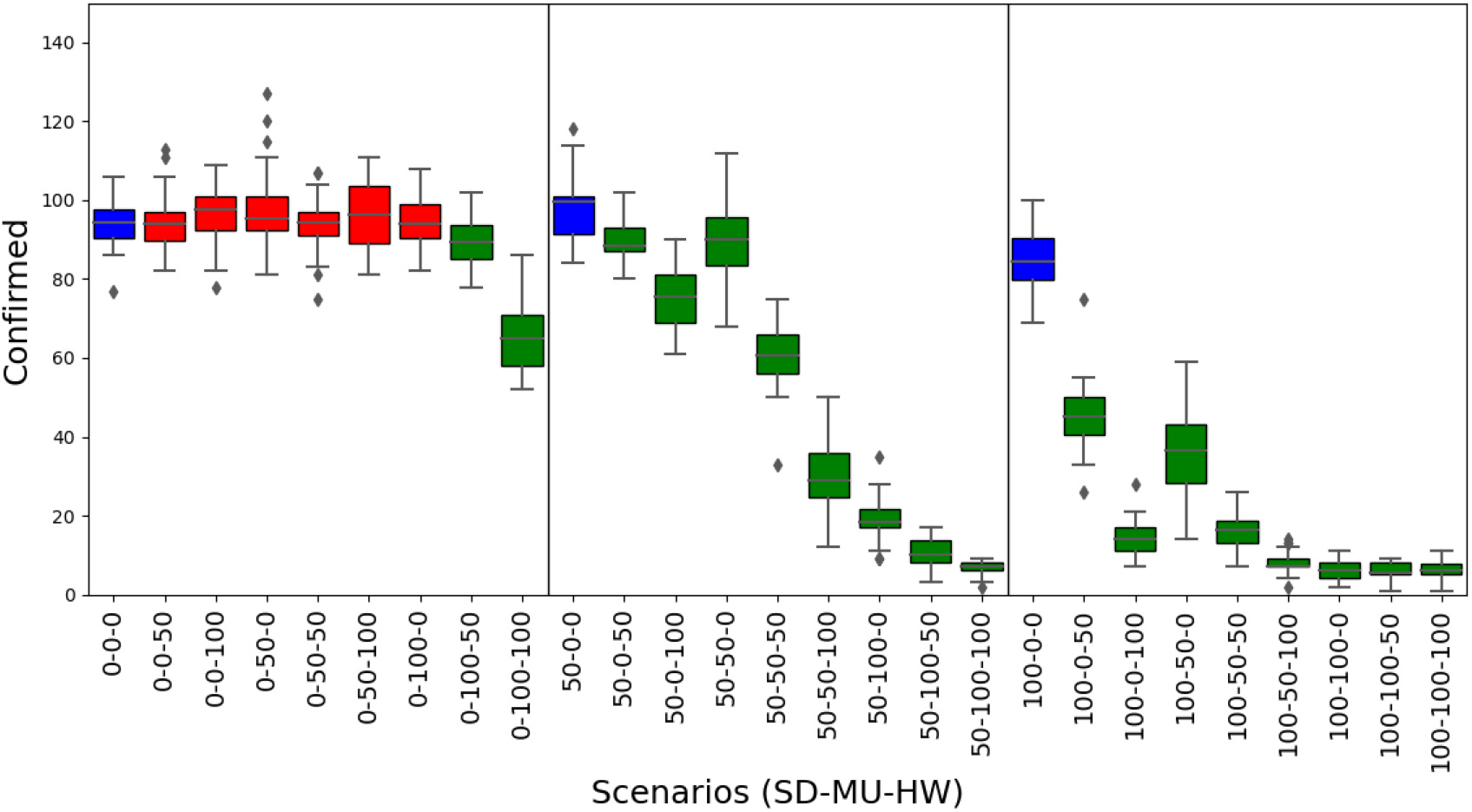
*Conirmed cases* results. Box plots represent average and standard deviation of the cumulative count of only those cases reported as diagnosed. See caption of Fig. 4 for interpretation details.

This behaviour is in line with the CFR overestimation of the IFR, mentioned before. For example, comparing the (0%, 0%, 0%) scenarios in Figure 5 vs Figure 6, these rates are IFR= 17% (68/400) vs CFR=72% (68/95); another example is the (100%, 0%, 0%) where these rates are IFR=12% (47/400) vs CFR=55% (47/85).

### Recovered

The behaviour of this epidemic indicator across all the simulated scenarios is shown in Figure 7. Here again, similar patterns appeared. In the left panel (SD=0%), only the (0%, 100%, 50%) and (0%, 100%, 100%) scenarios show a significant decrease of recovered agents compared to the baseline scenario, dropping from around 340, to 300 and to 200 recovered agents, respectively. Moreover, in the middle (SD=50%) and right panels (SD=100%), significant reductions are seen within all the scenarios compared to the baselines, with even sharper falls in the scenarios where half or more of the population adopt any or both of the other two measures (MU and HW), achieving values as fewer as approximately 20 agents (i.e 5% of the population). These reductions in the number of recovered agents emerge as a consequence of the mitigation or suppressing effects of these personal protection habits, implying much lower infections occurred, as the SIRE curves of Figure 2 and Figure 3 show evidently.

**Figure 7.**
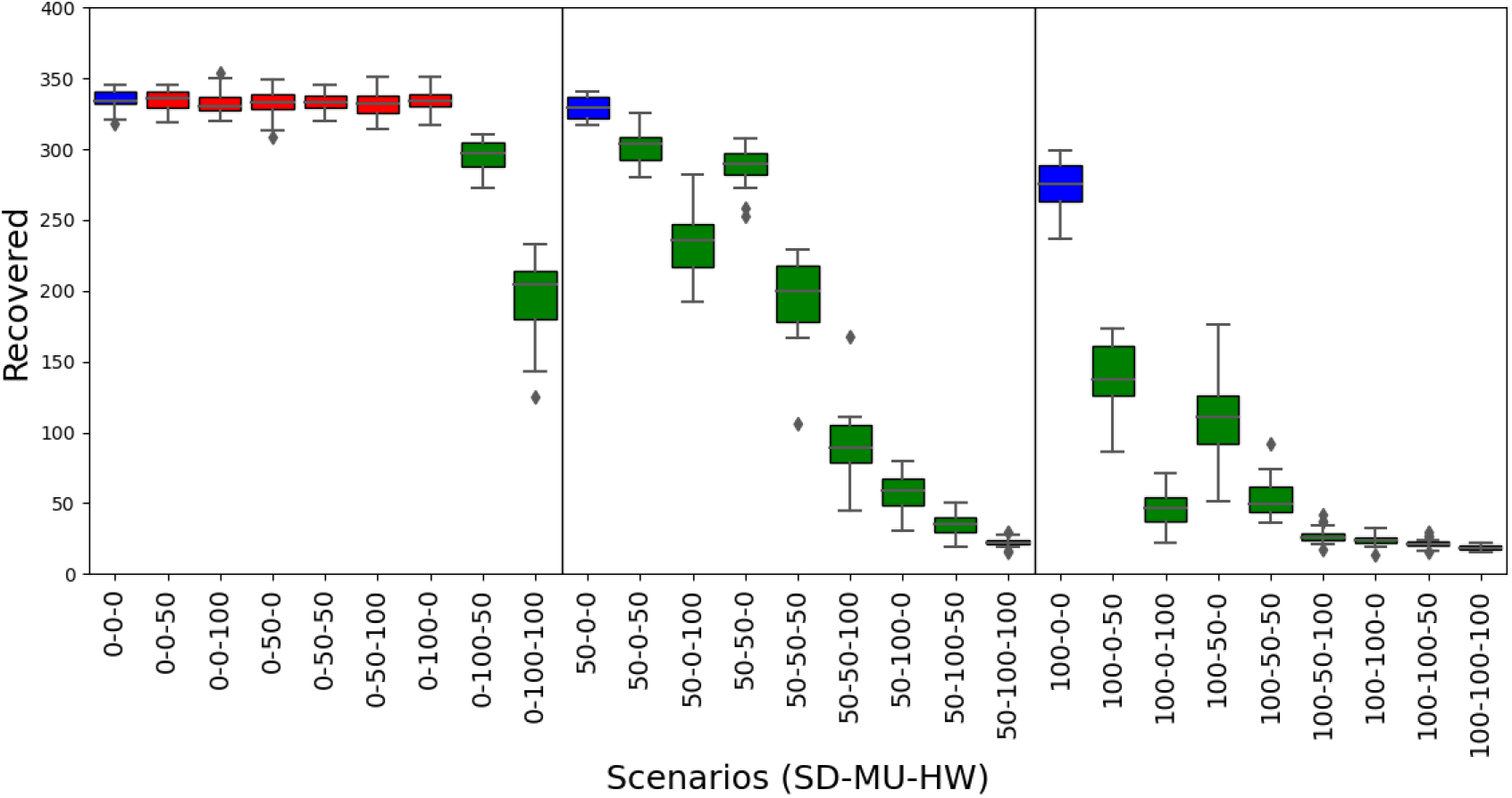
*Recovered* results. Box plots represent average and standard deviation of the cumulative count of agents that recovered from disease. See caption of Fig. 4 for interpretation details.

## Conclusion

As control policies are gradually lifted, the risk of a novel surge of the COVID-19 health crisis is highly likely, although it would be modulated by the adherence to voluntary individual protection habits by the community. We have illustrated the interplay between maintaining physical distance, using facial masks and washing hands regularly, with an agent-based simulation model of the spread of the epidemic on an artificial population. Despite the necessary assumptions made to simplify the model, our findings indicate that nearly-universal adoption of a single of these three habits alone, would not suffice to mitigate the rise of the infectious curve to controlled levels. Maintaining physical distance among persons plays a dominant role in decreasing the risk. However, in order to effectively suppress the growth of second outbreaks, the adoption by the population of one or both of the other two habits to some extent, is needed.

Some of the idealistic scenarios of universal adoption of these personal habits (100% scenarios), would be far to achieve in reality, due to many factors related with personal biases, socio-economic conditions and collective idiosyncrasies. Therefore in realistic scenarios, public health policies should be deployed to accompany these habit adoption, focusing on sustained mass– testing campaigns, rigorous contact tracing and mandatory household quarantine of confirmed cases (***Aleta et al., 2020***), so as to reduce or keep under control the spread of the disease in forthcoming epidemic waves during the “new normal”. Great attention should be taken by authorities to communicate clearly the evidence, uncertainties, risks and particularities of these personal protection strategies, without creating the sense of false dilemmas (***Escandón et al., 2020***) whilst aiming to ensure continued adherence of the community to these measures (***Thompson et al., 2020***).

There are interesting points not captured by the model, that we regard as worthy of a closer look. For example, effectiveness of material of face masks (it has been suggested that N95 respirators might be more strongly associated with protection from viral transmission than surgical masks or single-layer or fabric masks (***Chu et al., 2020***)). Additionally, considerations related to the adverse effects of universal masking wearing, including discomfort, irritation or misuse, psychological impact (***Bakhit et al., 2020***) or incidence in people with disabilities (inability of lip-reading for the hearing-impaired persons, difficulties with breathing for individuals with asthma or other respiratory disabilities (***Kohek et al., 2020; Chodosh et al., 2020***)), inadequate manipulation or risk of suffocation in preschool children (***Esposito and Principi, 2020***), and also environmental costs (***Allison et al., 2020; Klemeš et al., 2020***) might be taken as well into account. Besides, it is possible for masking habits to provoke misleading feelings of security causing to overlook other personal precautions in the community, although recent studies have found no association between mask usage and distance transgression (***Seres et al., 2020b***) or beyond that, have found evidence of individual attitudes to observe larger distance on waiting lines from someone wearing a face mask than from an unmask person, in a community with no face masking mandate (***Seres et al., 2020a***). The question of how elaborated an agent-based model should be to account for additional realistic features of the SARS-CoV-2 contagion phenomena, including infections via contaminated surfaces, population structure, spatial stratification, contact networks, host vulnerabilities and other sources of heterogeneity between individuals, pose interesting challenges from a computational modelling perspective (***Thompson et al., 2020***). Regarding the current version of our model, we are also interested in studying design mechanisms allowing agents to develop willingness to uptake the personal age protection habits as an emergent consequence of changes in individual biases or beliefs, affected by fluctuations in the popular consciousness.

## Data Availability

The simulation model has been posted and is available at the Modeling Commons public repository.

http://modelingcommons.org/browse/one_model/6423

## Supplementary material

1. **Mortality hypothesis tests for differences**. Heatmaps represent counts and *p*-values obtained by the Mann-Whitney U statistical test for permutations (SD%, MU%, HW%) with varying proportions of social physical distance, mask using and hand washing adoption by the population. Blue: baseline scenario, red: accept, green: reject.

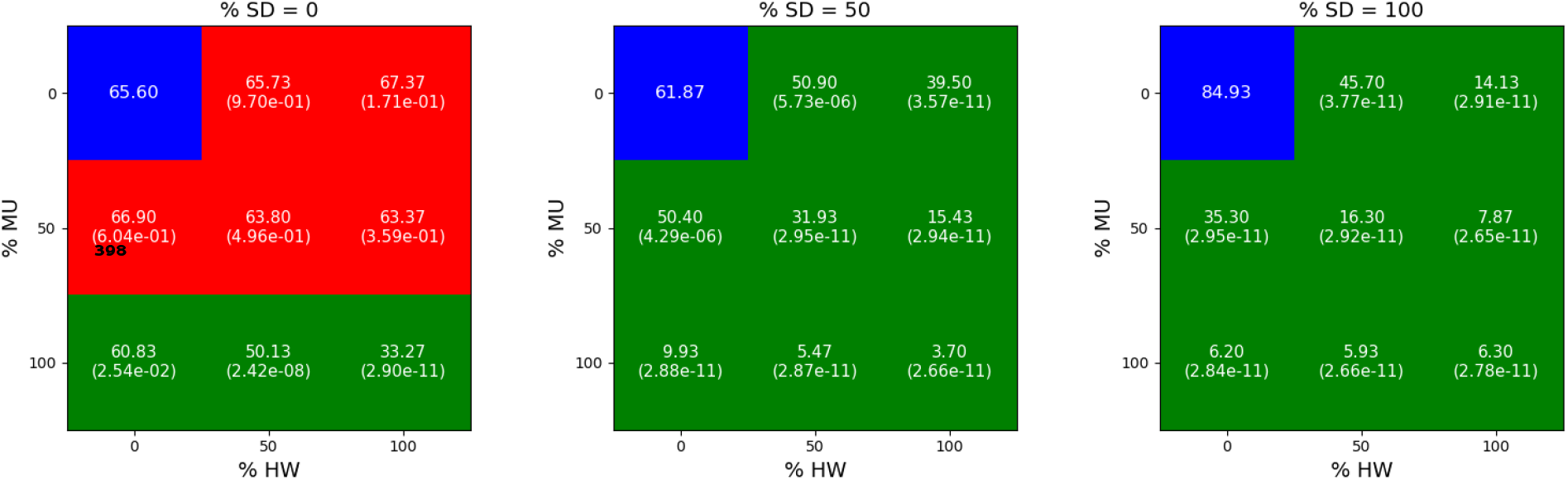
2. **Cases hypothesis tests for differences**. Heatmaps represent counts and *p*-values obtained by the Mann-Whitney U statistical test for permutations (SD%, MU%, HW%) with varying proportions of social physical distance, mask using and hand washing adoption by the population. Blue: baseline scenario, red: accept, green: reject.

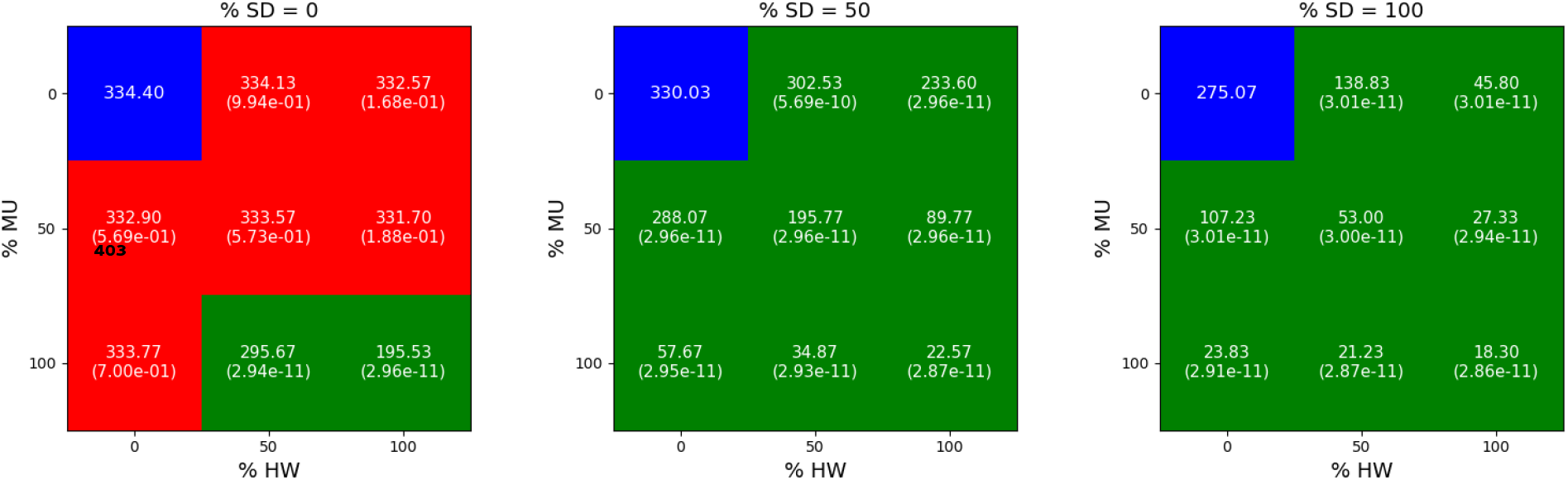
3. **Confirmed cases hypothesis tests for differences**. Heatmaps represent counts and *p*-values obtained by the Mann-Whitney U statistical test for permutations (SD%, MU%, HW%) with varying proportions of social physical distance, mask using and hand washing adoption by the population. Blue: baseline scenario, red: accept, green: reject.

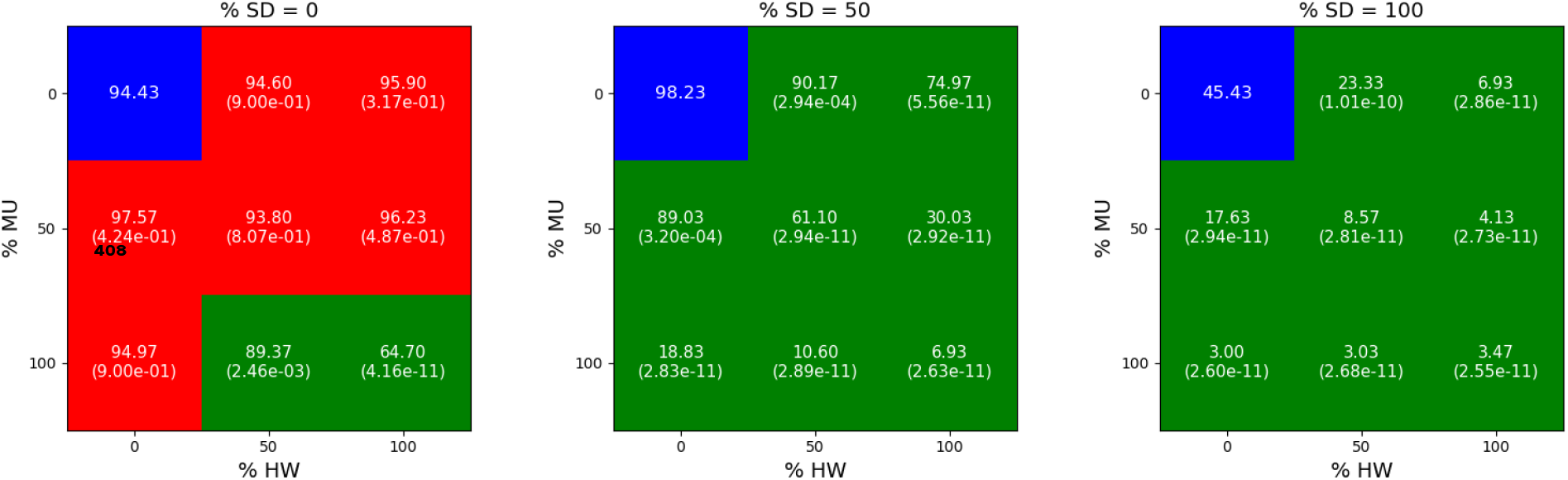
4. **Recovered hypothesis tests for differences**. Heatmaps represent counts and *p*-values obtained by the Mann-Whitney U statistical test for permutations (SD%, MU%, HW%) with varying proportions of social physical distance, mask using and hand washing adoption by the population. Blue: baseline scenario, red: accept, green: reject.

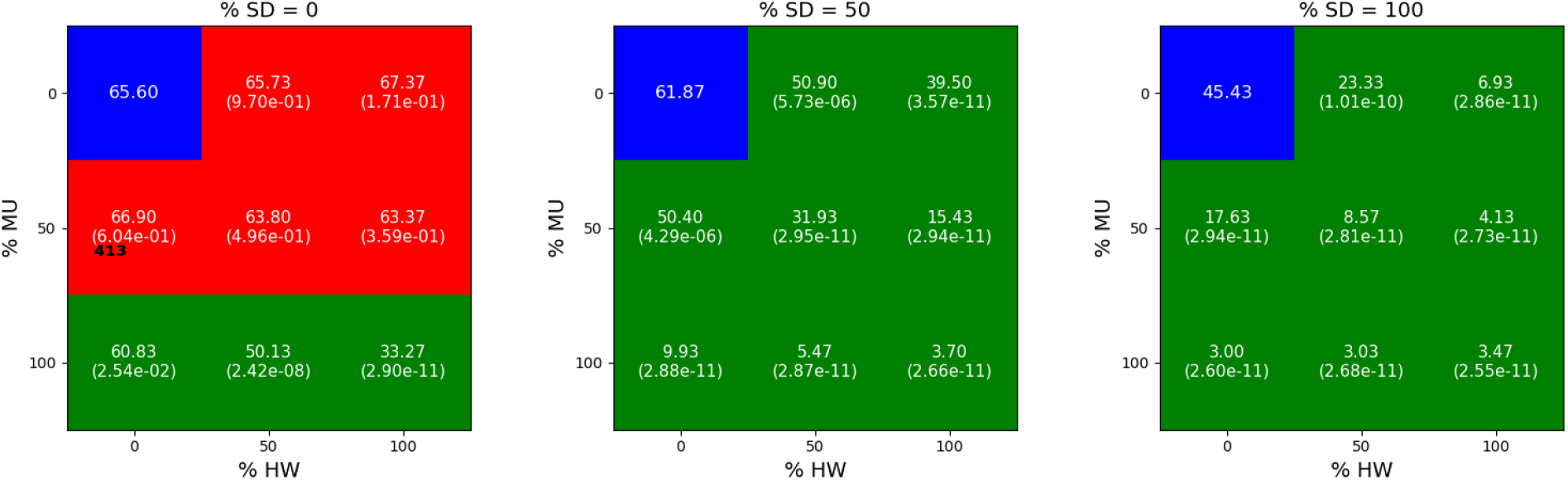

